# A clinical pilot study for personalized risk-based breast cancer screening utilizing the polygenic risk score

**DOI:** 10.64898/2026.03.07.26347839

**Authors:** Tone Hovda, Siim Sober, Peeter Padrik, Krista Kruuv-Kao, Eli M. Grindedal, Tone B. Aaman Vamre, Eline Eikeland, Solveig Hofvind, Kristine Kleivi Sahlberg

## Abstract

**Background:** Population-based mammographic screening is primarily age-based. However, breast cancer risk is multifactorial, and women may benefit from personalized risk-based screening. This pilot study aimed to explore the use of polygenic risk score (PRS) as a tool for risk stratification in personalized screening.

**Methods:** We included 80 women aged 40-49 years referred for clinical mammography. Exclusion criteria were prior breast cancer or premalignant breast disease, and previous genetic testing. After DNA collection, PRS was calculated from 2805 Single Nucleotide Polymorphisms (SNPs). Screening recommendations were based on each participant’s relative 10-year breast cancer risk estimated from PRS and compared with the 10-year risk of an average woman of the same age. Women with a self-reported family history of cancer meeting standard criteria were referred for gene panel testing for pathogenic variants in high-risk genes. A follow up questionnaire regarding participants’ experiences was distributed 6-9 months after PRS testing.

**Results:** Mean age was 45.2 years (SD 2.8). Mean relative 10-year breast cancer risk was 1.18 (SD 0.57). Based on PRS, 40 participants were recommended standard biennial screening 50-69 years, while 40 were advised to begin biennial screening before age 50. Among these, 7 were recommended annual mammography from when their 10-year risk reached twice that of an average 50-year-old. Twenty-one women underwent gene panel testing; no pathogenic variants in breast cancer genes were identified. Five women were advised annual mammography from 40-60 years due to family history of breast cancer, regardless of PRS. Most respondents viewed breast cancer risk assessment positively and did not report increased anxiety after testing.

**Conclusions:** Polygenic risk score testing may influence current screening recommendations and contribute to more personalized risk-based breast cancer screening strategies.

## 1. Introduction

Breast cancer is the most common cancer among women in Norway and worldwide (1, 2). Health authorities recommend mammography screening to reduce mortality from the disease through early detection (3, 4). Following European Commission guidelines (3), BreastScreen Norway invites all women aged 50-69 years to biennial mammography screening (5).

Breast cancer development is multifactorial, with risk factors including age, hormonal and reproductive history, mammographic density, lifestyle, environmental exposures, family history of cancer and genetic predisposition (4, 6, 7). Despite this complexity, population-based breast cancer screening programs typically apply a “one size fits all” approach, using age as the sole criterion, without addressing individual risk profiles (8–10). Although mammography screening has proved to reduce breast cancer mortality among participants, challenges persist, such as false positives, overdiagnosis, and diagnosis of advanced-stage cancers or interval cancers diagnosed between screening rounds (11–15). A more targeted and personalized screening strategy could enhance benefits and reduce harms for the individual women (9, 10).

Approximately one third of breast cancer cases are linked to genetic factors (16, 17), but only 4-5% are attributable to pathogenic variants in high-risk breast cancer genes, primarily *BRCA1* and *BRCA2* (17). In Norway, women with a family history of breast or ovarian cancer may be referred for genetic counselling and gene panel testing according to national guidelines (18). Genome-wide association studies have identified numerous low-risk genetic variations, single nucleotide polymorphisms (SNPs), which collectively explain about one-third of genetic susceptibility (7, 17, 19–22). While individual SNPs confer minimal risk, they can be combined into a polygenic risk score (PRS) expressing the total breast cancer risk attributed to these genetic variations. Modeling studies have explored PRS-based stratified screening alone, and combined with other risk factors, such as mammographic density and family history, as well as its integration into existing risk prediction models (7, 10, 23). Currently, PRS is not part of clinical prevention guidelines or systematic mammographic screening in Norway.

To inform future prospective studies and to address knowledge gaps, we conducted a clinical pilot study assessing PRS as a tool for personalized breast cancer screening among women aged 40-49 (pre-screening age) in Norway. Our objectives were to examine associations between PRS, mammographic density, family history, and high-risk gene analyses, and to explore women’s attitudes toward PRS-testing for personalized breast cancer screening.

## 2. Methods

The study was approved by the Norwegian Regional Committee for Medical Research Ethics (REK 494936). All participants provided written informed consent to study participation and use of collected data. The study adhered to the Declaration of Helsinki and relevant regulations. The trial is registered at ClinicalTrials.gov (NCT05731453, registration date 16/02/2023).

### 2.1 Study population

Women aged 40-49 referred to Vestre Viken breast center for clinical mammography due to breast symptoms between October 1 2022 and March 31 2023 were eligible. Exclusion criteria included current or prior breast cancer or premalignant breast disease, or previous genetic testing for familial breast cancer. Eligible women received an SMS after their visit with a link to study information and a digital consent form (Fig 1). Consenting participants completed an online questionnaire on life-style factors and family cancer history. Mammographic density was classified from clinical mammograms using the American College of Radiology’s Breast Imaging – Reporting and Data System (BI-RADS) classification (5^th^ edition), category a-d (24). Participants were scheduled for DNA sampling at the breast center. As a pilot study, enrollment was limited to 80 women. Data were managed using Ledidi Core (Ledidi AS, Norway).

**Fig 1.**
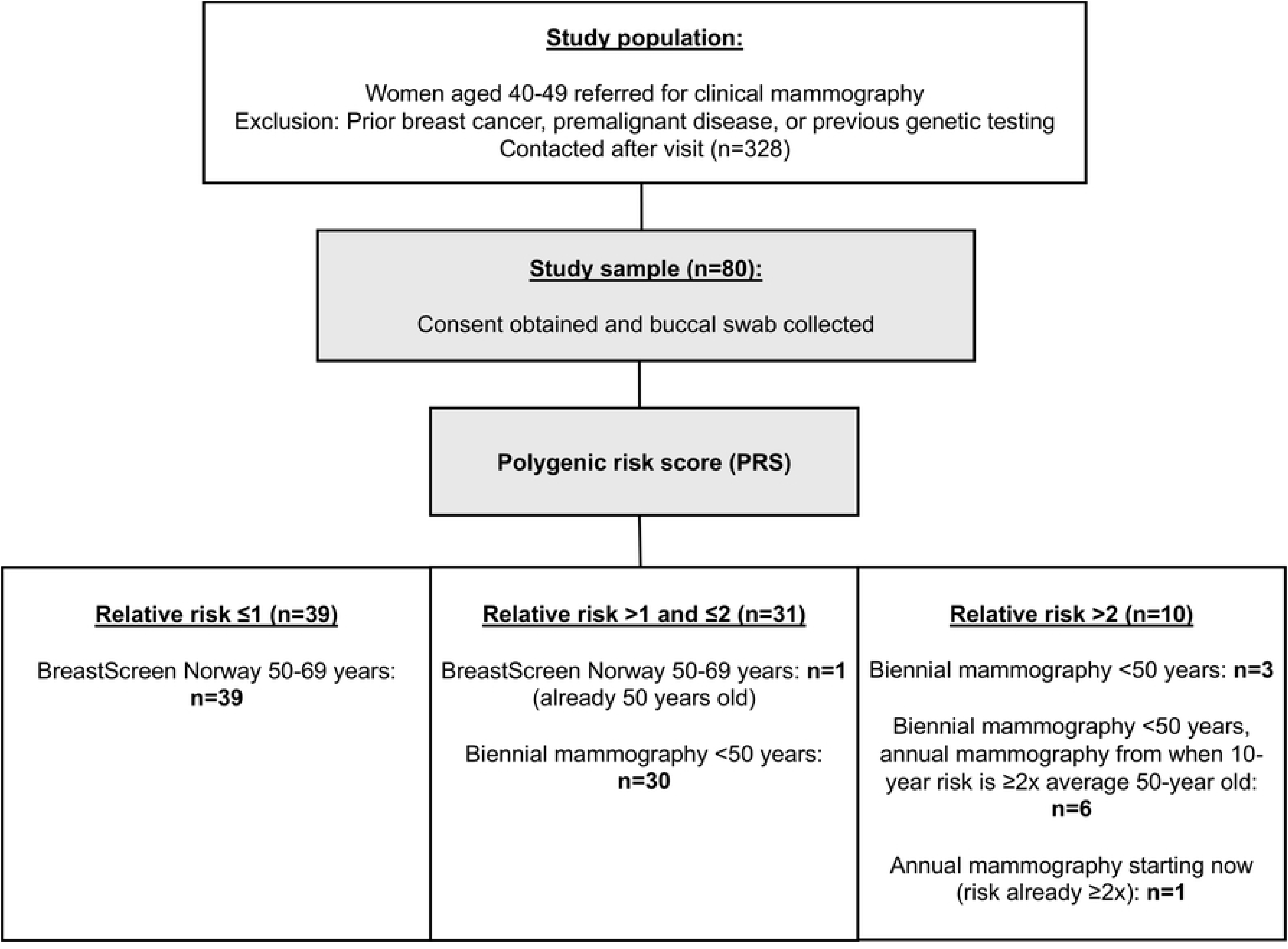
Flow chart.

### 2.2 Polygenic Risk Score (PRS) and AnteBC test

DNA was collected via Isohelix buccal swabs, stored in Isohelix Buccal Fix tubes (Isohelix, Cell Projects Ltd, UK) and sent to Antegenes (Tartu, Estonia) for PRS analysis using the CE-marked AnteBC test. This test, developed by Antegenes, estimates individualized breast cancer risk using PRS and has been validated in a Norwegian population (22). It analyzes 2803 breast cancer-associated SNPs to generate an individual PRS value and a 10-year breast cancer risk estimate. Test development was based on large datasets from two population biobanks, the Estonian Biobank (Estonian Genome Center, University of Tartu) and the UK Biobank. Detailed methodological descriptions of AnteBC and its computational framework are presented in Paadrik et.al. (25). Relative risk for breast cancer was calculated as each participant’s 10-year risk divided by the average 10-year risk for a Norwegian woman of the same age.

### 2.3 PRS-based screening recommendations

Screening recommendations were tailored to PRS results:

- Relative risk ≤1: Standard biennial mammographic screening at age 50-69 (Breast Screen Norway).
- Relative risk >1 and ≤2: Biennial screening beginning at the age when a woman’s 10-year risk reaches that of an average 50-year-old woman.
- Relative risk >2: Biennial screening from the age when the 10-year risk equals that of an average 50-year-old woman, and/or annual screening once the risk reaches twice that level.

Results and recommendations were communicated by postal letter, with the option of additional consultation if needed.

### 2.4 Family history of cancer

Participants reported cancer history among first- and second-degree relatives, not restricted to breast or ovarian cancer, using an online questionnaire. Those meeting national criteria for hereditary cancer testing (18) were referred to the Section for Hereditary Cancer at the Oslo University Hospital for genetic counselling and testing using a standard 29-gene panel *(APC, ATM, BAP1, BMPR1A, BRCA1, BRCA2, BRIP1, CDKN2A, CDK4, CHEK2, EPCAM, FLCN, HOXB13, MLH1, MSH2, MSH6, MUTYH, PALB2, PMS2, POLE, POLD1, PTEN, RAD51C, RAD51D, SDHB, SMAD4, STK11, TP53* and *VHL)*. Women with negative results but meeting guideline criteria (Table 1) were advised annual mammography from age 40-60 (18). In case of conflicting recommendations, the most intensive schedule was advised.

**Table 1.** Criteria for annual mammography from ages 40-60 in women without pathogenic variants in high-risk breast cancer genes.

| Annual mammography is recommended when <u>any</u> of the following criteria are met: |
| --- |
| • One first-degree relative diagnosed with breast cancer at $\leq 40$ years* |
| • Two-first degree relatives with breast cancer, with a mean age at onset $\leq 55$ years |
| • Three close relatives with breast cancer, with a mean age at onset $\leq 60$ years |
| • Four close relatives with breast cancer, regardless of age at onset |

\*If the affected relative was < 40 years at diagnosis, screening is recommended from age 30.

### 2.5 Follow-up

A digital questionnaire was sent to all participants 6-9 months post-inclusion to assess their experiences (S1 Table). Responses were recorded on a five-point scale and collapsed into three categories: disagree, neutral, and agree.

### 2.6 Statistical analyses

Descriptive statistics were used. Age, PRS-values, and relative 10-year risk were summarized as means with standard deviations (SD) and medians with interquartile ranges (IQR). Relative risk was categorized (≤1, >1 and ≤2, >2; ≤1.5 and>1.5). Analyses were stratified by mammographic density (low - BI-RADS a or b; high – BI-RADS c or d), genetic testing eligibility, and screening recommendations based on family history. Group differences were tested using two-sample t-tests, chi-square test, or Fisher’s exact test; p<0.05 was considered statistically significant. Results from the follow-up questionnaire about women’s experiences were presented as proportions (disagree, neutral, agree) for all and stratified by relative risk (≤1 or >1). All analyses were performed in IBM SPSS Statistics v.29.

## 3. Results

A total of 320 eligible women were invited to achieve the target sample size of 80, as specified in the study protocol. This resulted in a 25% response rate (80/320). The mean age of participants was 45.2 years (SD 2.8), and the median age was 45.0 years (IQR 43.0-47.5) (Table 2).

**Table 2.** Age, family history of cancer, PRS-value, and relative PRS-based breast cancer risk stratified by mammographic density.

|  | Total | BI-RADS a/b | BI-RADS c/d | p-value |
| --- | --- | --- | --- | --- |
| **N (%)** | 80 (100%) | 41 (51%) | 39 (49%) |  |
| **Age (years)** |  |  |  |  |
| Mean (SD) | 45.2 (2.8) | 44.4 (2.9) | 46.0 (2.6) | 0.009 |
| Median (IQR) | 45.0 (43.0-47.5) | 44.0 (42.0-47.0) | 47.0 (44.0-48.0) |  |
| **Family cancer history** |  |  |  |  |
| Positive | 25 (31%) | 12 (29%) | 13 (33%) | 0.70 |
| Negative | 55 (69%) | 29 (71%) | 26 (67%) |  |
| **PRS-value** |  |  |  |  |
| Mean (SD) | 0.13 (1.09) | 0.23 (1.01) | 0.02 (1.17) | 0.38 |
| Median (IQR) | 0.15 (-0.52-0.88) | 0.34 (-0.44-0.88) | -0.15 (-0.79-0.60) |  |
| **Relative risk** |  |  |  |  |
| Mean (SD) | 1.18 (0.57) | 1.22 (0.53) | 1.14 (0.61) | 0.58 |
| Median (IQR) | 1.07 (0.80-1.48) | 1.16 (0.82-1.48) | 0.94 (0.71-1.30) |  |
| **Relative risk** |  |  |  |  |
| ≤1 | 39 (49%) | 18 (44%) | 21 (54%) | 0.34 |
| >1 and ≤2 | 31 (39%) | 19 (46%) | 12 (31%) |  |
| >2 | 10 (12%) | 4 (10%) | 6 (15%) |  |
| **Relative risk** |  |  |  |  |
| ≤1.5 | 61 (76%) | 31 (76%) | 30 (77%) | 0.89 |
| >1.5 | 19 (24%) | 10 (24%) | 9 (23%) |  |

### 3.1 Polygenic risk score and screening recommendations

The mean PRS value was 0.13 (SD 1.09) and the median PRS value was 0.15 (IQR -0.52-0.88). Mean relative 10-year breast cancer risk based on PRS for participants compared to age-matched population was 1.18 (SD 0.57), median 1.07 (IQR 0.80-1.48) (Table 2).

Among the 80 women included in the study, 49% (39/80) had a relative breast cancer risk of 1 or lower based on their PRS. Thirty-nine percent (31/80) had a relative risk between 1 and 2, while 12% (10/80) had a relative risk greater than 2 (Table 2).

Based on these results, 50% (40/80) were advised standard biennial mammographic screening at ages 50-69. Further, 49% (39/80) were advised to start biennial screening before age 50, at the age when their 10-year risk equaled that of an average 50-year-old. Among these, six women (15%) were further recommended annual screening once their risk doubled that of an average 50-year-old. One of the 80 participants (1%) was advised to begin annual screening immediately because her risk was already at that level (Fig 1).

When dichotomized at an alternative threshold of 1.5, 76% (61/80) were classified as low risk (≤1.5) and 24% (19/80) as high risk (>1.5) (Table 2), although this stratification was not linked to separate recommendations.

Regarding breast density, 51% (41/80) were categorized as BI-RADS a or b (low density), and 49% (39/80) as BI-RADS c or d (high density). No statistically significant association was observed between PRS and mammographic density (Table 2).

### 3.2 Family history and genetic testing

Based on family history, 29% (23/80) were referred for genetic counselling and testing with a 29-gene-panel; 70% (16/23) had a family history of breast or ovarian cancer. Two participants did not provide samples, leaving 21 tested participants. No pathogenic variants in breast cancer genes were identified, although one participant carried a *CDKN2A* variant associated with melanoma and pancreatic cancer risk. Five women (24%) met criteria for annual mammography from age 40-60 based on family history (Table 1) (18).

Mean PRS among gene panel tested participants was 0.45 (SD 1.01) versus 0.03 (SD 1.10) for non-tested (p=0.13). Median PRS-values were 0.39 (IQR -0.31-1.18) for tested and -0.05 (IQR -0.69-0.73) for non-tested (Table 3). A statistically significantly higher proportion of tested women had a relative breast cancer risk >1.5 compared with non-tested (43% (9/21) versus 18% (10/57), p=0.021) (Table 3). For the five women recommended annual screening from age 40-60 based on family history, mean PRS was 1.39 (SD 0.30) compared with 0.15 (SD 0.99) for others (p=0.018). Further, all five also had relative risk >1, reinforcing intensive screening recommendations (Table 3).

**Table 3.** PRS value and relative risk stratified by family history of cancer.

| Genetic counselling and gene panel testing due to positive family history of cancer* |  |  |  |
| --- | --- | --- | --- |
|  | Yes | No | p-value |
| N (%) | 21 (27%) | 57 (73%) |  |
| PRS-value |  |  |  |
| Mean (SD) | 0.45 (1.01) | 0.03 (1.10) | 0.13 |
| Median (IQR) | 0.39 (-0.31-1.18) | -0.05 (-0.69-0.73) |  |
| **Relative risk** |  |  |  |
| ≤1 | 8 (38%) | 29 (51%) | 0.60 |
| >1 and ≤2 | 10 (48%) | 21 (37%) |  |
| >2 | 3 (14%) | 7 (12%) |  |
| **Relative risk** |  |  |  |
| ≤1.5 | 12 (57%) | 47 (82%) | 0.021 |
| >1.5 | 9 (43%) | 10 (18%) |  |
| **Recommended annual mammography screening based on family history of cancer** |  |  |  |
|  | **Yes** | **No** | **p-value** |
| **N (%)** | 5 (24%) | 16 (76%) |  |
| **PRS-value** |  |  |  |
| Mean (SD) | 1.39 (0.68) | 0.15 (0.99) | 0.018 |
| Median (IQR) | 1.55 (1.18-1.73) | 0.09 (-0.45-1.02) |  |
| **Relative risk** |  |  |  |
| ≤1 | - | 8 (50%) | 0.059 |
| >1 and ≤2 | 3 (60%) | 7 (44%) |  |
| >2 | 2 (40%) | 1 (6%) |  |

\*Data not available: n=2

### 3.3 Women’s experiences

Of the 80 participants, 73% (58/80) responded to the follow-up questionnaire. Among respondents, 43% (25/58) had a PRS-based relative risk ≤1 and 57% (33/58) had a relative risk >1. In total, 24% (14/58) of the respondents were referred for gene panel testing due to family history.

Most women (98% (57/58)) found buccal swab sampling acceptable (Fig 2). Communication of PRS results was considered satisfactory by 79% (46/58), while 10% (6/58) were neutral. Another 10% (6/58) were dissatisfied, all of whom had a relative risk >1 (Fig 3), and they expressed a preference for phone communication rather than letters. Four women with increased risk contacted the breast center for additional information.

**Fig 2.**
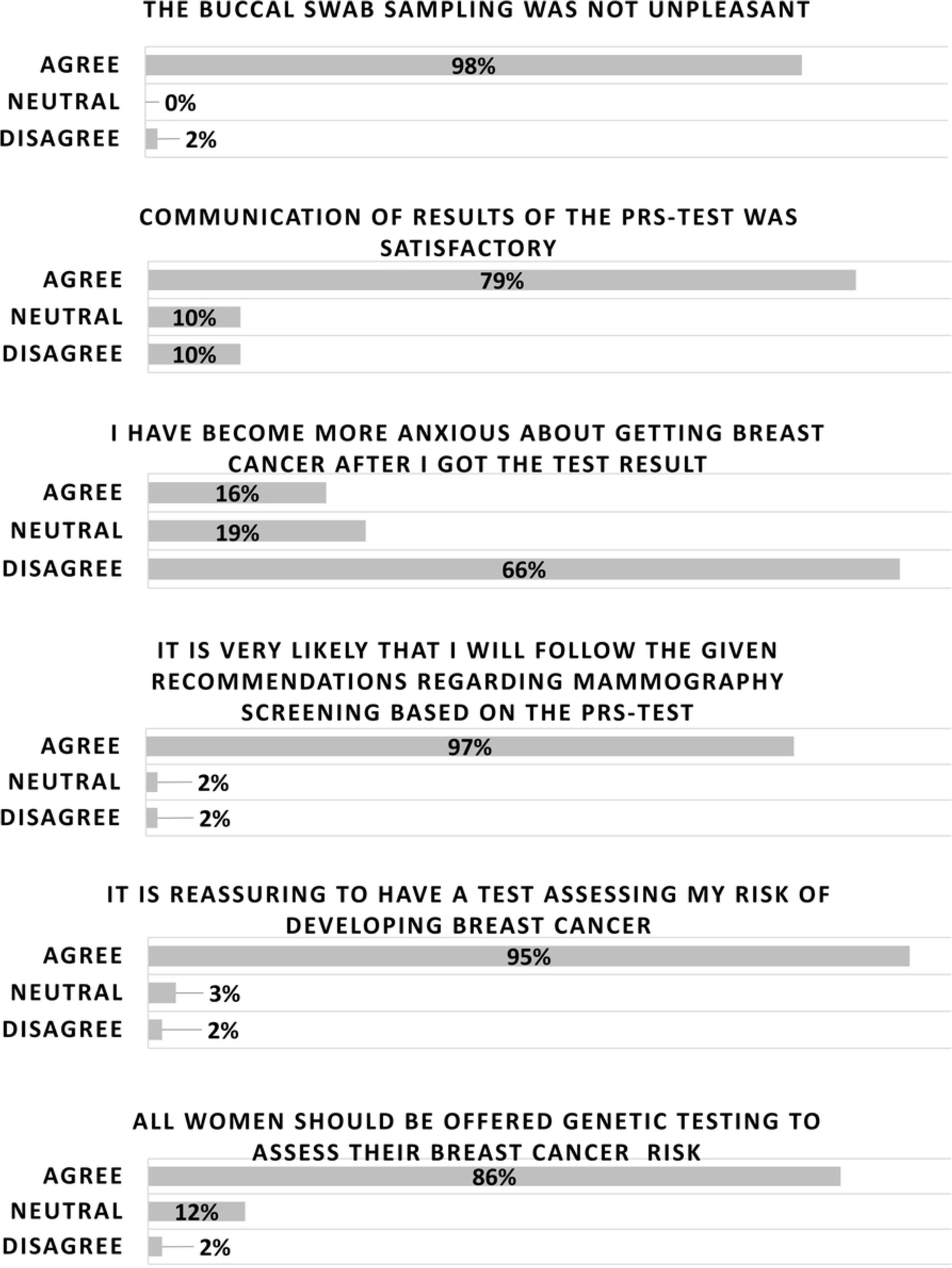
Women’s experiences with participation in the study. Total number of respondents: 58.

**Fig 3.**
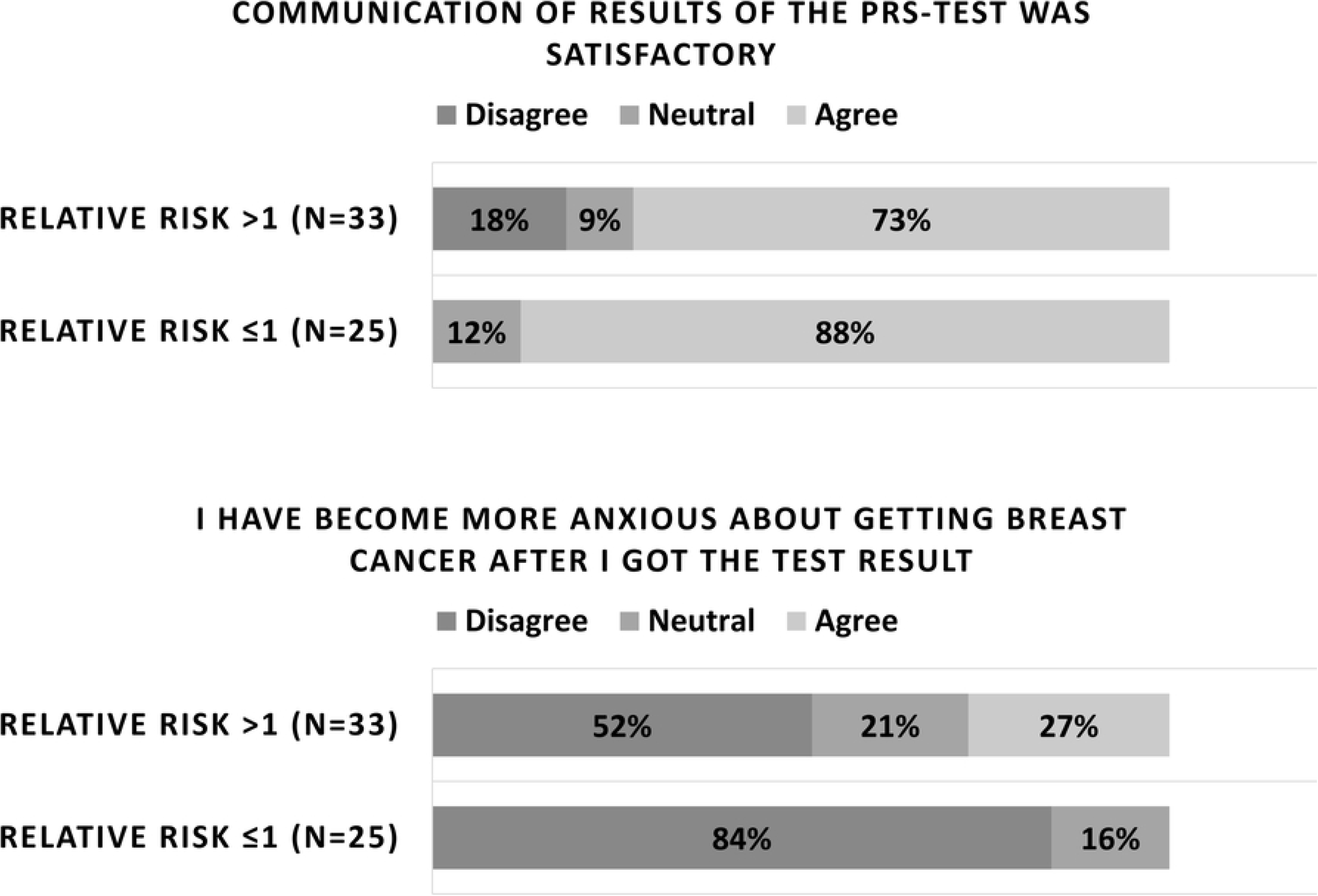
Women’s views on communication of results and anxiety related to breast cancer. Data are stratified by PRS-based relative risk (≤1 vs. >1). Total number of respondents: 58.

Regarding anxiety, 66% (38/58) of respondents disagreed that testing made them more anxious, 19% (11/58) were neutral, and 16% (9/58) agreed (Fig 2). All the respondents who were neutral or who reported increased anxiety had a relative risk >1 (Fig 3).

Almost all respondents, 97% (56/58), intended to follow the screening recommendations given, and 95% (55/58) found testing reassuring. Furthermore, 86% (50/58) agreed that all women should be offered genetic testing to assess breast cancer risk (Fig 2).

Among the 14 respondents referred to genetic counselling and gene panel testing due to family history, 86% (12/14) were satisfied with the process, 93% (13/14) intended to follow recommendations, and 86% (12/14) considered testing important for themselves and their families (Fig 4).

**Fig 4.**
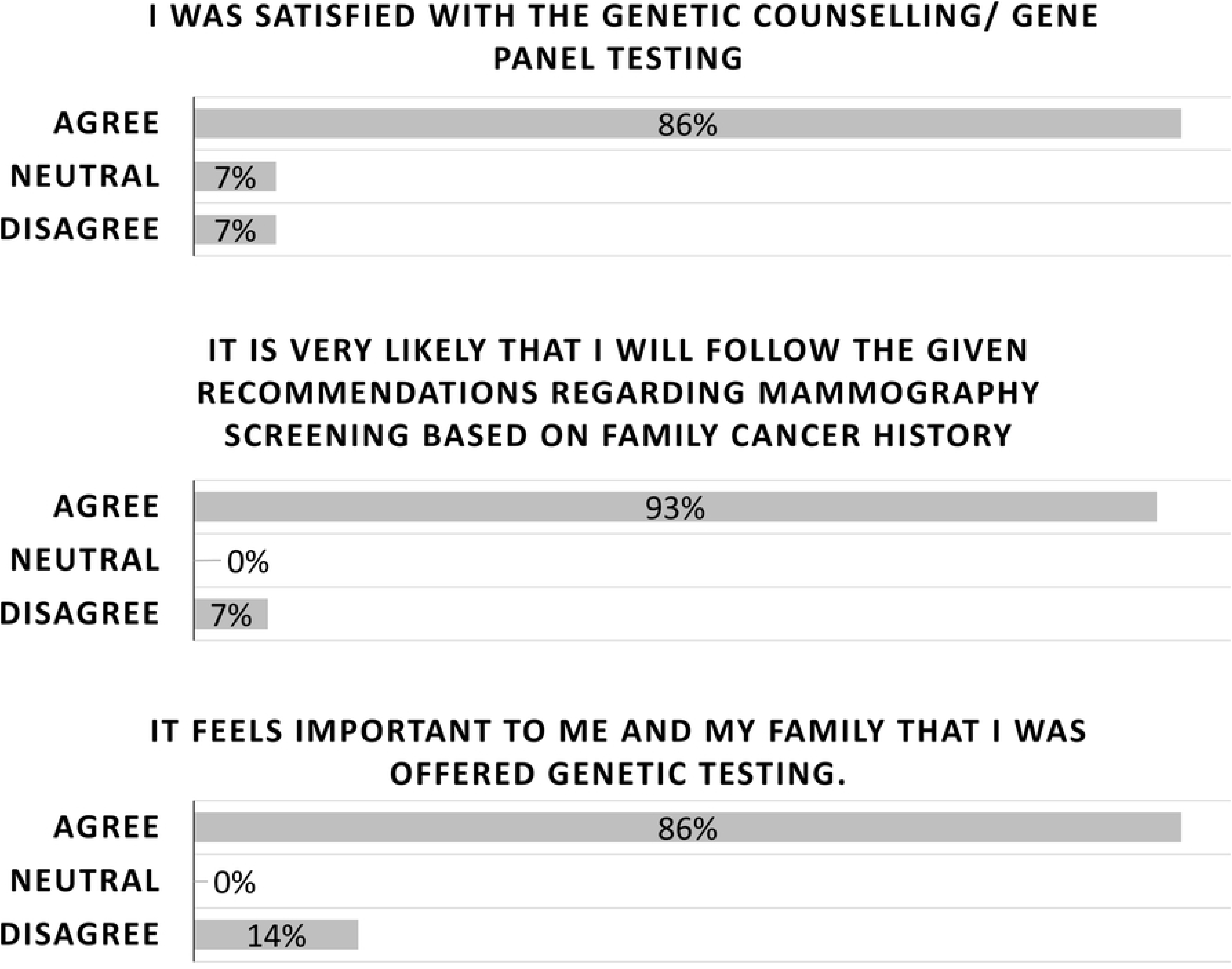
Women’s experiences following referral to genetic counselling and gene panel testing due to family cancer history. Total number of respondents: 14.

Respondents’ detailed answers to all questions are provided in S2 Table.

## 4. Discussion

We conducted a prospective clinical pilot study that included 80 women aged 40-49 years with no prior history of breast cancer or premalignant breast disease. We analyzed each participant’s polygenic risk score (PRS) and explored their experiences with study participation. Half of the women had a PRS-based 10-year breast cancer risk higher than average for women of the same age and were therefore advised to begin mammographic screening earlier and/or undergo screening more frequently than standard biennial screening. Based on self-reported family history, 27% underwent genetic counselling and gene panel testing for pathogenic variants in high-risk cancer genes in accordance with national guidelines. Participants were generally satisfied with their involvement in the study and expressed positive attitudes toward receiving an individualized breast cancer risk assessment. A minority reported increased breast cancer-related anxiety after receiving their test results, all of whom had an elevated PRS-based risk.

Currently, population-based mammographic screening is largely based on age as the sole risk factor. In BreastScreen Norway, women receive their first screening invitation at age 50 (more precisely between ages 48-52, depending on residence and the biennial screening interval), and each woman receives at total of ten invitations (5). However, as our results demonstrate, some women reach the breast cancer risk level of an average 50-year-old at considerably younger age and may therefore benefit from an earlier onset of mammographic screening. This pattern has also been shown by Akdeniz et al., who analyzed Norwegian data using the AnteBC 2803-SNP test. In their study, women in the 99^th^ PRS percentile had a lifetime cumulative breast cancer risk nearly six times higher than women in the 1^st^ percentile (22).

For some women, annual screening or the use of supplementary imaging techniques may be beneficial. Polygenic risk scores have shown strong value in risk-stratification models, particularly when combined with other established risk factors such as breast density and family history (17, 26). As previously noted, family history is already an accepted basis for risk-adapted screening in many countries, including Norway (18, 27). In our study, we did not observe a statistically significant association between PRS and breast density; however, the study was likely underpowered to detect clinically meaningful differences. These findings are consistent with previous research showing that PRS and breast density independently influence breast cancer risk without being correlated (28, 29).

Breast cancer risk is estimated to be 4-6 times higher in women with extremely dense breasts compared to those with predominantly fatty breasts (30). The European Society of Breast Imaging (EUSOBI) recommends that women with extremely dense breasts be offered breast MRI screening every 2-4 years (31). However, breast density is not routinely used for risk assessment or risk stratification in Norway, either within the national screening program or in clinical mammography practice.

Among women referred for gene panel testing, a statistically significantly higher proportion had a relative 10-year breast cancer risk >1.5 compared with those not referred, even though referral to gene panel testing was based on family history of all cancers, not solely breast cancer. In addition, women recommended more intensive screening due to family history of breast cancer had a statistically significantly higher mean PRS value. These findings support results from larger studies showing that common genetic variations (SNPs) contribute meaningfully to the hereditary component of breast cancer and may play a role in risk-based mammographic screening (16, 17, 19). Risk-based stratification, offering women at higher risk more intensive screening, has the potential to improve early detection and reduce mortality and morbidity for the individual women. However, earlier start and increased screening frequency also raise concerns about potential harm, such as false positives, overdiagnosis and overtreatment (10, 32). Moreover, increasing the number of screening examinations for a subset of women will inevitably increase workload and costs for the health care system, underscoring the need for careful evaluation of risk thresholds for intervention. In this clinical pilot, we used a relative risk >1 to recommend earlier start of mammographic screening, resulting in half of the participants being advised to start screening before age 50. Raising the threshold to relative risk >1.5 reduced the proportion in the increased-risk group to 24%. Incorporating PRS into risk assessment models including other risk factors can modify the risk assessment for individual women and may influence the number of additional screening examinations required. Risk prediction models such as the Breast and Ovarian Analysis of Disease Incidence and Carrier Estimation Algorithm (BOADICEA)/CanRisk are valuable tools in this context (7, 33), but were not applied in this study as they are not routinely used in Norway.

Another important dimension of personalized, risk-based breast cancer screening is the potential to downscale screening in women with low risk. Reducing screening intensity in this group might be considered more controversial than intensifying screening for women at high risk. Van Ravesteyn (32) examined the balance between benefits and harms across different screening intervals for low-risk women and demonstrated that both reduced breast cancer risk and low breast density substantially diminish the absolute benefit of screening. Consequently, the added value of biennial compared with triennial screening in these women appeared limited. Van den Broek (10) reported increased life-years gained, fewer breast cancer deaths, and the most favorable ratio of life-years gained to overdiagnosis when screening was tailored using a combination of family history and PRS. These findings are also highly relevant for cost-effectiveness considerations. Two large ongoing trials, My Personalized Breast Screening (MyPebs) trial (34) and the Women Informed to Screen Depending on Measures of Risk (WISDOM) trial (8) are currently evaluating whether risk-based screening is non-inferior to standard age-based screening with respect to rates of stage II cancers. They aim to include 85,000 and 100,000 women, respectively. Their results will provide essential evidence to inform future screening strategies. Implementation of personalized screening programs also requires careful attention to logistics, particularly when genetic testing is involved. In our study, all participants attended an appointment at the breast center for buccal swab sampling. For large-scale implementation, more efficient strategies such as mailing self-collection kits and using digital platforms for communicating test results and screening recommendations would be necessary. A solely age-based screening program is inherently equitable and well accepted, as all participants receive the same examination regardless of individual risk. Incorporating genetic information into screening recommendations inevitably raises ethical, legal, and social considerations, as women must make decisions that extend beyond attendance alone. It is anticipated that not all women will consent to genetic testing for breast cancer risk assessment. This may challenge principles of equality if those declining testing are assigned screening pathways that could be perceived as suboptimal. It is therefore essential to ensure that screening offered to women who do not consent to genetic testing remains at least equivalent to the current standard. In our study, participation was 25% among the 320 women invited, suggesting a degree of reluctance toward genetic testing and risk assessment in the general population. Although most respondents expressed support for offering all women an assessment of their genetic breast cancer risk, this finding is biased by the fact that all respondents had already agreed to take part in a study with genetic testing as a core component. As one participant reflected: *“I was not sure if it was a good idea to test. I was worried to know if the increased risk would hurt me more than help.”* Among respondents with increased risk based on PRS, approximately one in four reported heightened anxiety about breast cancer following participation, meaning three in four did not. Some degree of anxiety is an expected and reasonable response to the disclosure of elevated risk. Structured communication and follow-up for these women are therefore important, extending beyond the provision of screening recommendations alone. This principle is also embedded in the MyPeBS study protocol (34). Understanding the impact of positive test results on women’s quality of life, including short- and long-term psychological responses, remains a key area for future research.

The most prominent limitation of our study was the sample size of only 80 participants, which likely rendered it underpowered to detect meaningful statistical differences for some variables. However, as a pilot study, the primary objective was to inform the design and feasibility of a future, larger prospective study. Furthermore, because all women eligible for inclusion had been referred for clinical mammography at a breast center, the study population may not be representative for women in their pre-screening age decade and may inherently differ in baseline breast cancer risk. This limits the external generalizability of our findings. The exclusion of women with a prior history of breast cancer, and those already referred for genetic testing due to family history, may also have shifted the study cohort toward a lower-risk group compared with the standard population. Selection bias was likely, as participation required an active choice both to enroll in the study and to complete the follow-up survey. Women with negative attitudes toward genetic testing or risk-stratified screening may have been less likely to participate. This concern is, however, relevant for future implementation as well, as PRS-based screening cannot realistically become mandatory within population-based screening programs. Finally, some of the questions in the follow-up questionnaire may be perceived as somewhat leading. Nonetheless, respondents showed broad agreement across most questions, suggesting that more neutral phrasing would be unlikely to alter the overarching patterns of responses.

## 5. Conclusion

Polygenic risk scores have the potential to support a transition from the current uniform breast cancer screening model to a more personalized risk-based approach. PRS may also provide valuable risk assessment for women outside organized screening programs. An optimal implementation strategy will likely require the integration of PRS with established clinical risk factors, including family history and mammographic density. Furthermore, advances in artificial intelligence-based image analysis are expected to play an increasingly significant role in future risk stratification efforts. Ensuring clear communication of results and appropriate follow-up will be essential to maintain acceptability and trust among women invited to participate in risk-adapted screening pathways. To guide implementation, larger prospective studies are needed, including comprehensive evaluations of benefits and harms, feasibility, acceptability, and cost-effectiveness.

## Data Availability

Patient level data cannot be shared publicly because these are human research participant data and contain potentially identifying or sensitive patient information. Aggregated data from the patient survey have been included in the Supporting information files.

## Supporting information

**S1 Table. Follow-up questionnaire sent to all participants 6-9 months after PRS-testing.** Response was given for each question on a five-point scale: Completely disagree, slightly disagree, neutral, slightly agree and totally agree.

**S2 Table. Participants’ answers to all questions in the follow-up questionnaire as listed in S1 Table**. Response given on a five-point scale. Total respondents question no. 1-8: n=58. Total respondents question no. 9-13: n=14.

